# Explainable advanced electrocardiography has a high negative predictive value for ruling out significant coronary artery disease on cardiovascular computed tomography

**DOI:** 10.1101/2025.03.02.25323182

**Authors:** Manoj Rajamohan, Daniel Loewenstein, Maren Maanja, Kevin Yang, Chosita Cheepvasarach, Todd T Schlegel, Martin Ugander, Rebecca Kozor

## Abstract

**BACKGROUND:** Advanced electrocardiography (A-ECG) has been used to improve the diagnostic performance of the ECG in a number of cardiac disease states. We hypothesised that A-ECG can improve the diagnostic assessment of intermediate risk chest pain by optimising an A-ECG score for significant coronary artery disease (CAD) by cardiovascular computed tomography (CCT).

**METHODS:** Participants attending an outpatient rapid access chest pain clinic underwent a 12-lead ECG and CCT. Significant CAD was defined as luminal stenosis >50%. Multivariable logistic regression was performed using measures from the conventional ECG, derived vectorcardiography, and singular value decomposition measures of waveform complexity.

**RESULTS:** Of included patients (n=171, 60% male, age 59±13 years), 37 (22%) had >50% stenosis in at least one coronary artery, with single, double, or triple vessel disease in 38%, 38%, and 24%, respectively. A four parameter A-ECG score to detect significant CAD had an area under the receiver operating characteristic curve [95% confidence interval] of 0.87 [0.78–0.94], sensitivity 89 [69–97]%, specificity 82 [68-94]%, positive predictive value 55 [43–78]%, negative predictive value 96 [92–99]%, positive likelihood ratio 4.6 [2.9–13.1] and inverse negative likelihood ratio 6.4 [2.9–27.2].

**CONCLUSION:** A-ECG can rule out significant CAD on CCT with a high negative predictive value and overall good diagnostic performance. This supports the use of A-ECG to screen patients in a chest pain clinic setting who would benefit from further testing or not.

## Introduction

Chest pain is one of the most common presentations to emergency departments with significant cost associated with admission to hospital for further diagnostic assessment and treatment (1). By obviating the need for hospitalisation, the rapid access chest pain clinic (RACPC) model has been shown to be a valuable outpatient clinical pathway for the safe and efficient assessment, diagnosis and treatment of patients presenting with symptoms suspicious of cardiac chest pain and at low to intermediate risk(2, 3). A RACPC seeks to reduce the financial burden associated with unnecessary hospital admission for management of patients with low to intermediate risk chest pain, through the use of risk stratification algorithms such as the HEART(4) score and highly accurate diagnostic tests including cardiovascular computed tomography (CCT)(5).

The conventional 12-lead ECG is the initial diagnostic test used in the risk stratification of chest pain. However, ECG has a relatively low sensitivity and specificity for the diagnosis of clinically significant coronary artery disease outside of the diagnosis of myocardial infarction (6, 7). The use and interpretation of the ECG has been largely unchanged in the past few decades, however recent improvements in digital technology, specifically in signal processing and biomedical computing, have enabled newer innovative techniques of ECG analysis which have demonstrated significant promise, collectively termed advanced electrocardiography (A-ECG).

A-ECG refers to the integration of conventional ECG parameters with other electrocardiographic analysis techniques including singular value decomposition measures of waveform complexity and vectorcardiographic (VCG) measures, all derived from the resting 12-lead ECG. In clinical practice, A-ECG analysis can be performed on digitally acquired ECG files, both rapidly and inexpensively (8).

We hypothesised that A-ECG can improve the diagnostic assessment of intermediate risk chest pain in the RACPC. The aim of our study was to create an A- ECG score for significant coronary artery disease (CAD) as detected by CCT in a RACPC patient population.

## Methods

### Patient population

All patients attending an outpatient RACPC at Royal North Shore Hospital (Sydney, Australia) between February 2017 and January 2020 who underwent digital electrocardiography and cardiovascular computed tomography (CCT) were included. The study was approved by the local ethics committee (Northern Sydney Local Health District Human Research Ethics Committee) and all participants provided written informed consent or a waiver of consent was obtained. Exclusion criteria were prior diagnosis of CAD, prior myocardial infarction and previous revascularisation, including percutaneous coronary intervention and coronary bypass grafting.

### ECG acquisition and analysis

Resting 12-lead ECG data for each subject were collected from the local ECG storage system and exported into xml files. A-ECG semi-automatic software developed in-house was used to analyse the xml files, and this encompassed conventional ECG measures of durations and amplitudes, derived VCG measures, and singular value decomposition measures of waveform complexity. Further exclusion criteria included non-sinus rhythm, bundle branch block (QRS>120ms), pre-excitation, heart rate > 100bpm, and corrupted or uninterpretable ECG files.

### CCT analysis

Each clinically acquired CCT was analysed by a study investigator (MR) blinded to the A-ECG results for the presence, distribution, and severity of CAD. Significant CAD was defined as luminal stenosis >50% in any coronary artery.

### Statistical Analysis

Statistical analysis was performed in R (R Foundation for Statistical Computing, version 4.1.2, Vienna, Austria). Continuous variables are reported as mean±SD and count data as number and frequency. Baseline characteristics were compared using the t-test, or Mann-Whitney U-test where values not normally distributed. The A-ECG score was derived using stepwise forward logistic regression of conventional and advanced ECG measures. After fixing the parameters and coefficients following logistic regression, bootstrap resampling was performed 2000 times. The reported area under the receiver operating characteristics curve (AUC) and other performance metrics including sensitivity, specificity, negative and positive predictive values, and positive and inverse negative likelihood ratios are median values generated from the bootstrapped iterations and 95% confidence intervals. The A- ECG score that yielded the highest area under the receiver operator curve was chosen. The Youden index identified optimized cut-offs for sensitivity and specificity. A minimum of ten events per incremental A-ECG parameter was accepted for the scores.

## Results

The final analysis included 171 patients (60% male, age 59±13 years). Table 1 summarises the baseline patient characteristics. Of these, 37 (22%) had >50% stenosis in at least one coronary artery. The total cohort was deemed low-to-intermediate risk for major cardiac events as indicated by a HEART score of 3±1 (range 1-6). The group with at least one significant stenosis had a higher risk score compared to the group with no significant stenosis (HEART score 4±1 vs 3±1; p<0.001). The significant stenosis group was older (67±12 vs 56±13 years, p<0.001) and had a higher proportion of males (84% vs 54%, p<0.001). They were also more likely to be hyperlipidaemic (68% vs 39%, p=0.002) and smokers (59% vs 28%, p<0.001). Rates of hypertension and diabetes did not between the two groups, and both groups were overweight as indicated by body mass index (BMI) (27±5 vs 28±7 kg/m2, p=0.3). Of the patients with a significant stenosis, the incidence of single, double, or triple vessel disease was 38% (14/37), 38% (14/37) and 24% (9/37), respectively. There were no instances of significant left main disease.

**Table 1:** Baseline patient characteristics.

| Parameters | All | <50% stenosis | >50% stenosis | p-value |
| --- | --- | --- | --- | --- |
| Number of patients, n (%) | 171 (100) | 134 (78) | 37 (22) |  |
| Age, years | $59 \pm 13$ | $56 \pm 13$ | $67 \pm 12$ | <0.001 |
| Male sex, n (%) | 103 (60) | 72 (54) | 31 (84) | <0.001 |
| HEART score | $3 \pm 1$ | $3 \pm 1$ | $4 \pm 1$ | <0.001 |
| Body mass index, kg/m<sup>2</sup> | $27 \pm 5$ | $27 \pm 5$ | $28 \pm 7$ | 0.32 |
| Hypertension, n (%) | 62 (36) | 46 (34) | 16 (43) | 0.32 |
| Diabetes, n (%) | 18 (11) | 15 (11) | 3 (8) | 0.59 |
| Smoker, n (%) | 60 (35) | 38 (28) | 22 (59) | <0.001 |
| Hyperlipidaemia, n (%) | 77 (45) | 52 (39) | 25 (68) | 0.002 |

Continuous variables reported as mean ± SD and count data as number and percent.

The four A-ECG features that had the strongest association with significant CAD were: (1) the frontal plane QRS loop axis at maximum voltage (degrees), (2) peak root mean square voltage of the QRS vector magnitude (mV), (3) natural logarithm of the R-wave duration in lead V6 [log(ms)], and (4) R wave amplitude in lead aVF (μV). Mean values for these A-ECG measures are included in Table 2.

**Table 2:**
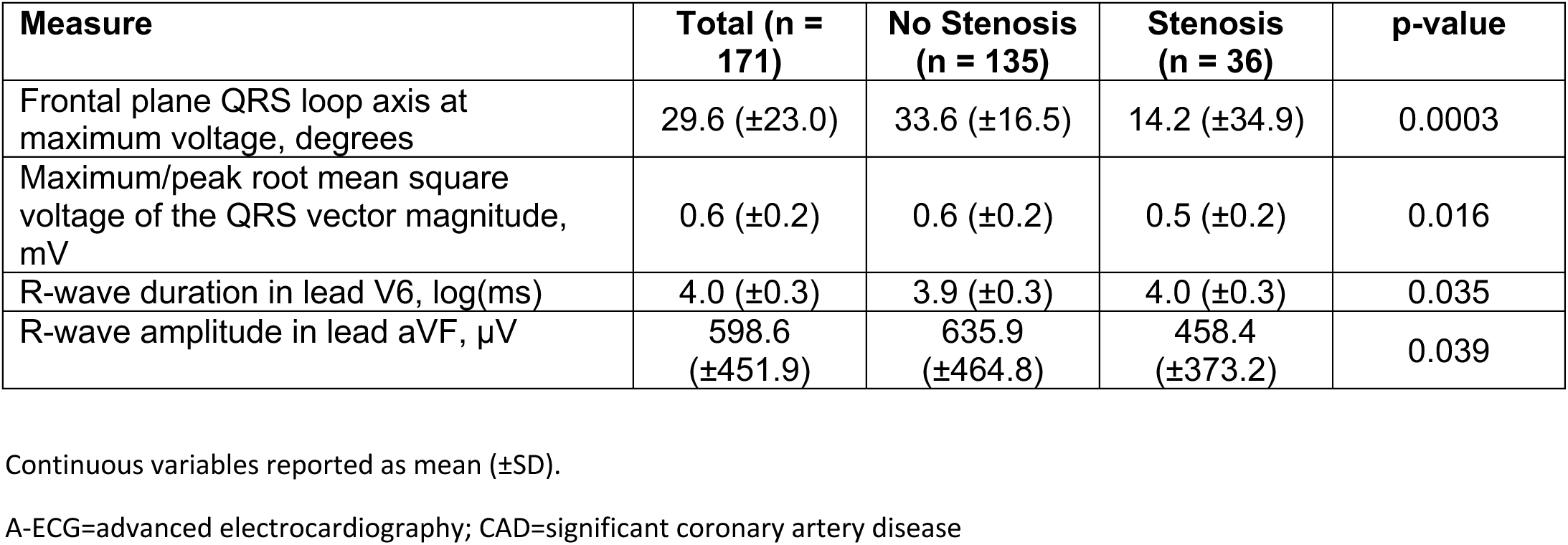
A-ECG measures in all patients, stratified by presence of significant CAD.

| Measure | Total (n = 171) | No Stenosis (n = 135) | Stenosis (n = 36) | p-value |
| --- | --- | --- | --- | --- |
| Frontal plane QRS loop axis at maximum voltage, degrees | 29.6 (±23.0) | 33.6 (±16.5) | 14.2 (±34.9) | 0.0003 |
| Maximum/peak root mean square voltage of the QRS vector magnitude, mV | 0.6 (±0.2) | 0.6 (±0.2) | 0.5 (±0.2) | 0.016 |
| R-wave duration in lead V6, log(ms) | 4.0 (±0.3) | 3.9 (±0.3) | 4.0 (±0.3) | 0.035 |
| R-wave amplitude in lead aVF, μV | 598.6 (±451.9) | 635.9 (±464.8) | 458.4 (±373.2) | 0.039 |

Continuous variables reported as mean ( ± SD).
A-ECG=advanced electrocardiography; CAD=significant coronary artery disease

These features were combined to derive a four parameter A-ECG score to detect significant CAD with an area under the receiver operating characteristic (ROC) curve [95% confidence interval] of 0.87 [0.78–0.94], sensitivity 89 [69–97]%, specificity 82 [68-94]%, positive predictive value 55 [43–78]%, negative predictive value 96 [92–99]%, positive likelihood ratio 4.6 [2.9–13.1] and inverse negative likelihood ratio 6.4 [2.9–27.2]. The measures included in the final four-parameter A-ECG score, and the intercept and coefficients for the regression equations are presented in Table 3. The ROC curve demonstrating diagnostic ability of the A-ECG score for detection of significant CAD is shown in Figure 1. A-ECG score ranking for the entire cohort is shown in Figure 2 with optimised cut-off score of >50% deemed high probability for significant CAD.

**Figure 1:**
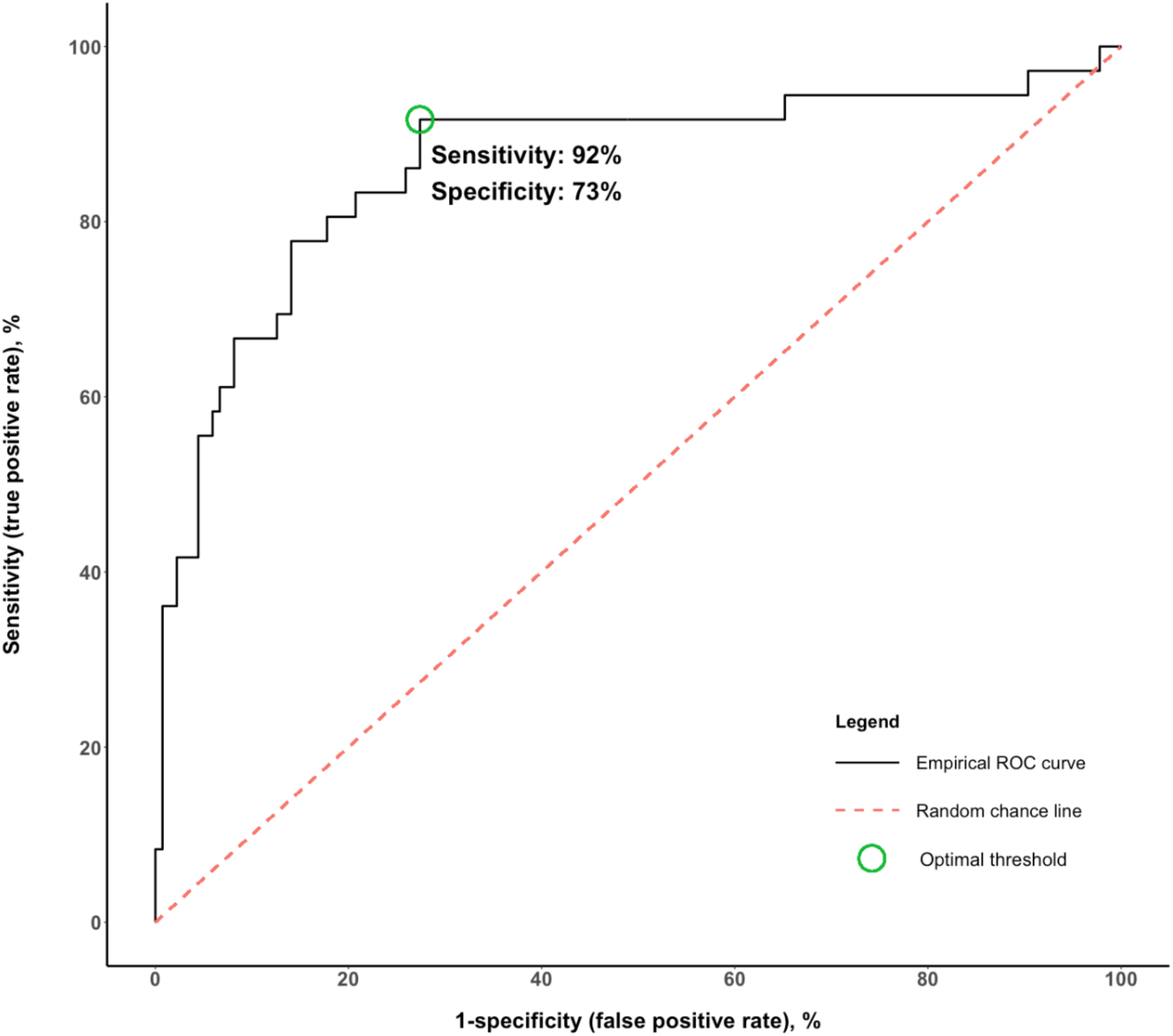
Receiver operating characteristic curve demonstrating diagnostic ability of the four parameter A-ECG score for detecting CAD. A-ECG=advanced electrocardiography; AUC=area under the curve; CAD=significant coronary artery disease; CI=confidence interval; INLR=inverse negative likelihood ratio; NPV=negative predictive value; PLR=positive likelihood ratio; PPV=positive predictive value; ROC=receiver operating characteristic; Sens=sensitivity; Spec=specificity

**Figure 2:**
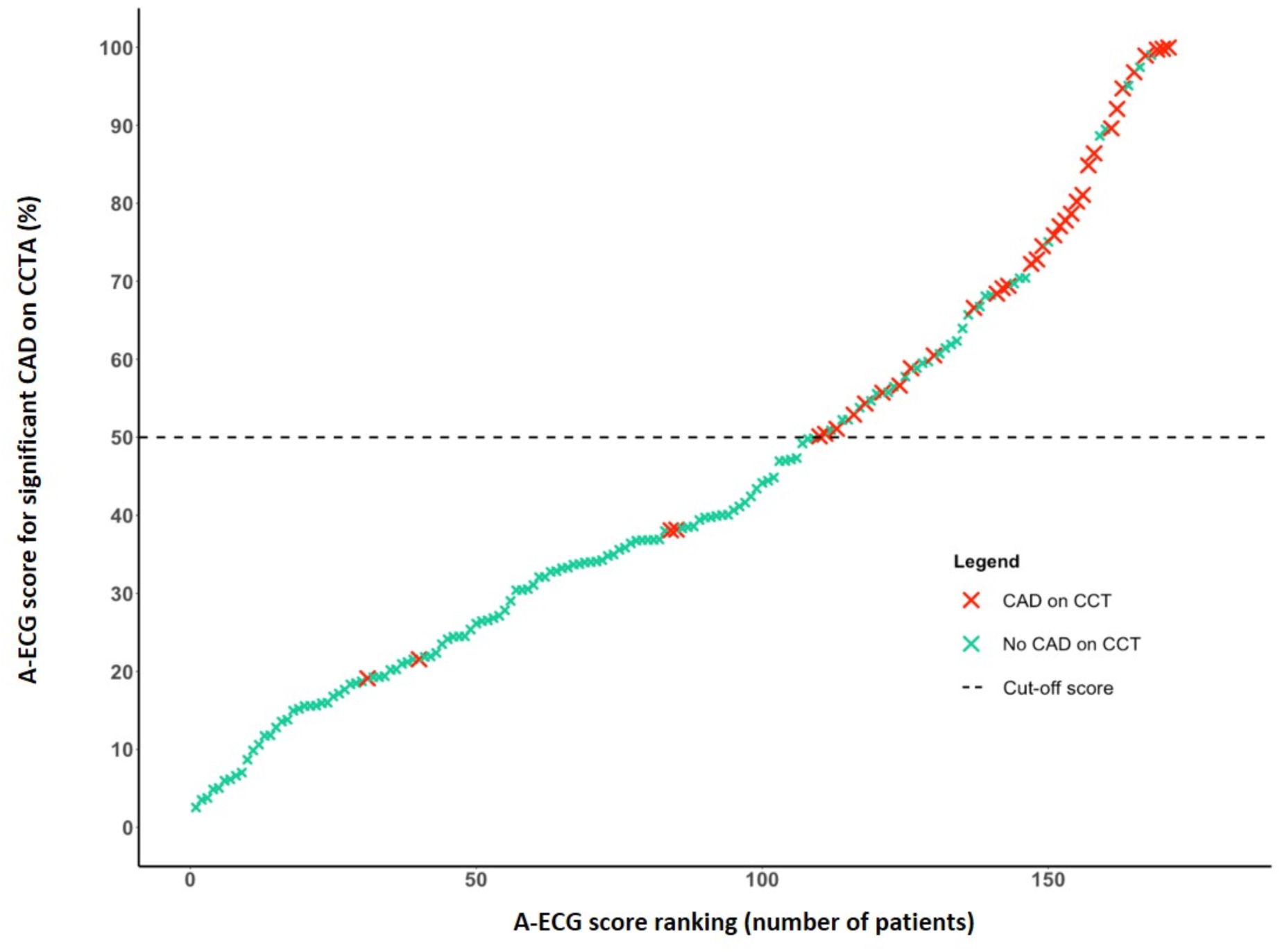
Scatter plot demonstrating A-ECG score ranking and cut-off score for significant CAD A-ECG=advanced electrocardiography; CAD=coronary artery disease; CCTA=cardiac computed tomography angiography

**Table 3:**
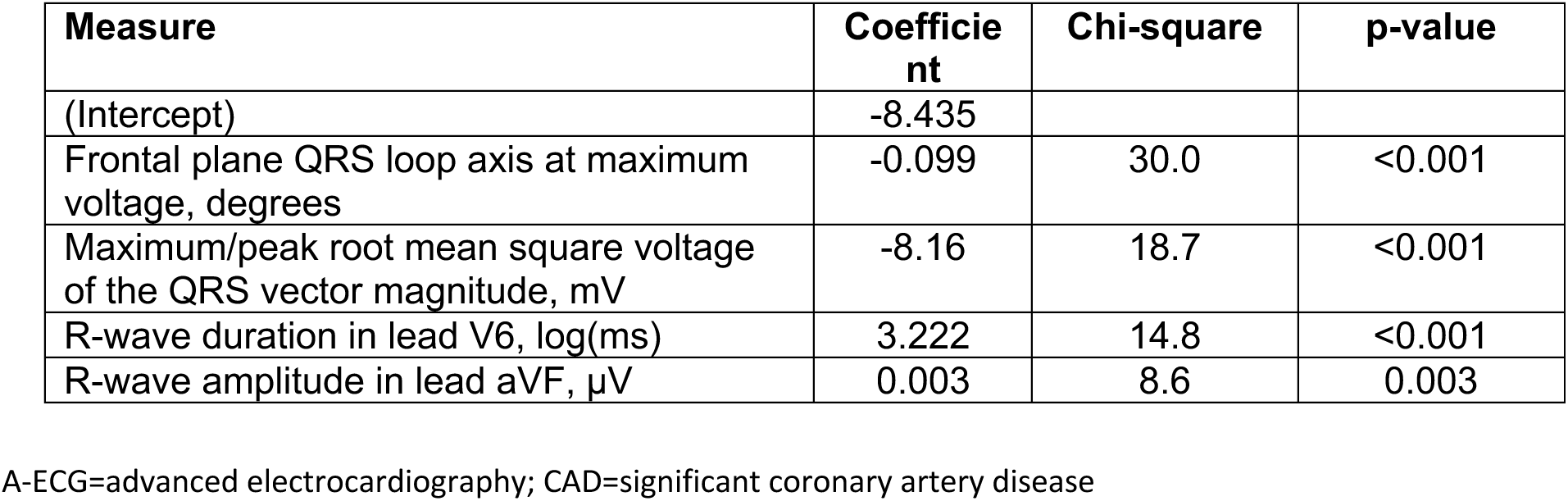
Measures included in the four parameter A-ECG score to detect CAD.

## Discussion

A-ECG can rule out significant CAD on CCT with a high negative predictive value and good overall diagnostic performance using four variables derived from the resting standard 12-lead ECG. This supports the use of A-ECG to screen patients in a chest pain clinic setting who would benefit from further testing or not.

This is the first study to investigate the use of A-ECG in the evaluation of CAD as detected by CCT in a low-to-intermediate risk population through a rapid access chest pain clinic. Prior studies have shown the ability of A-ECG to detect the presence of catherization-proven or other imaging proven CAD with higher sensitivity and specificity than the conventional ECG (8, 9). Additionally, other studies have demonstrated the utility of A-ECG in identifying left ventricular systolic dysfunction, left ventricular hypertrophy and other cardiac conditions (10–12). Whilst invasive coronary angiography remains the reference standard for the detection of CAD, the use of CCT in this study reflects the increased uptake of this imaging modality in RACPCs given its high sensitivity, specificity and negative predictive value and recent incorporation into clinical guidelines (13–15).

Low-intermediate risk (HEART score ≤ 6) chest pain patients, compared to their high risk counterparts (HEART score > 6), suffer a lower rate of major adverse cardiac events (MACE) at 30 days, 2-17% vs. 50% respectively(16). The assessment and management of these patients in RACPC has been shown to be a cost effective strategy(5), however the use of A-ECG as a rapid and inexpensive screening tool in this setting also shows promise. As illustrated in Figure 3, conventional ECG demonstrated no significant abnormalities in study patients A and B, however A-ECG analysis revealed a high probability for significant coronary stenosis in the latter. Subsequent CCT revealed a high grade stenosis in the mid right coronary artery for patient B.

**Figure 3:**
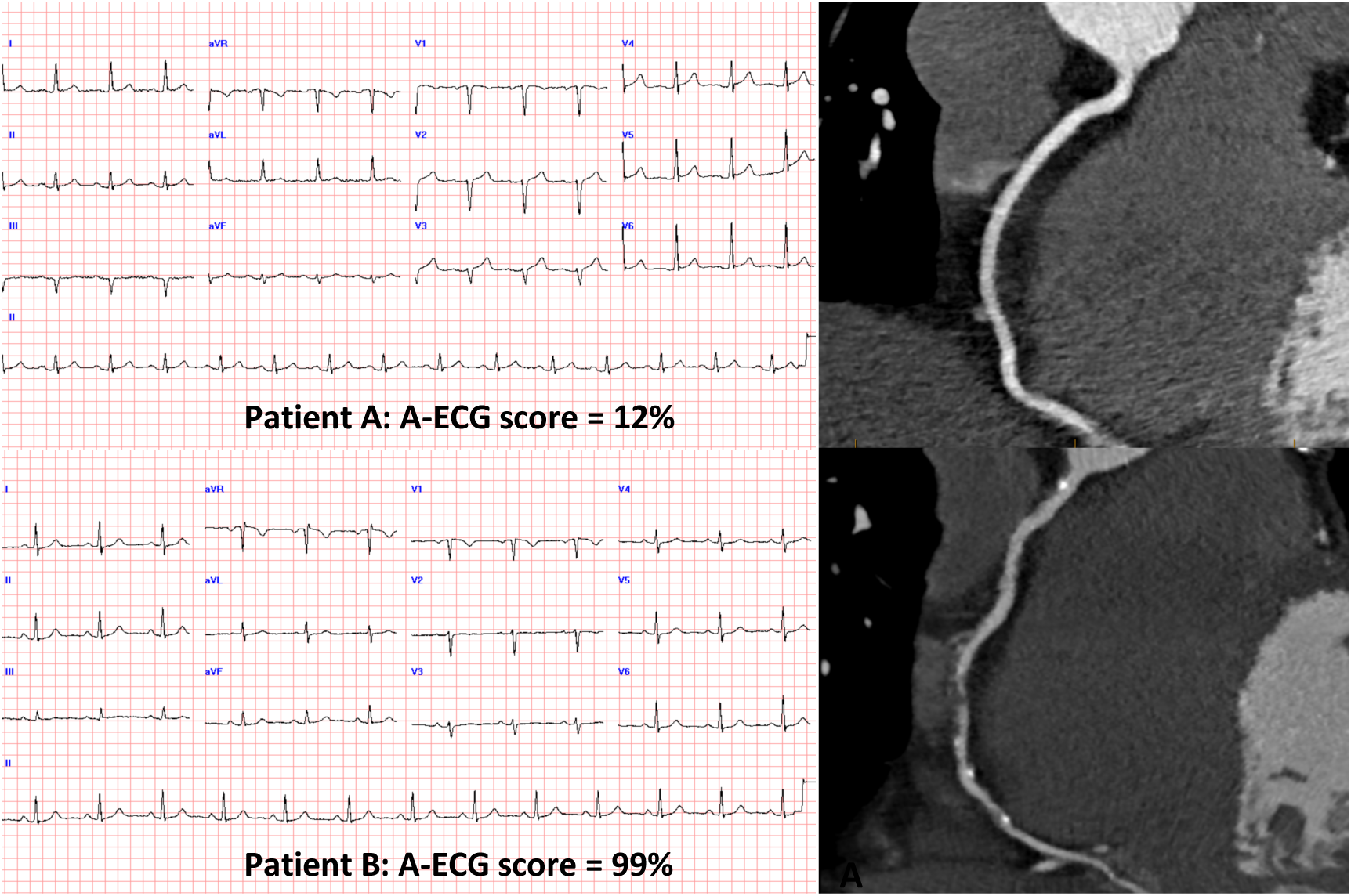
The top panel shows a visually unremarkable ECG for Patient A with low probability A-ECG score for having significant coronary artery disease (12%), and cardiac computed tomography angiography for same patient showing a normal right coronary artery (top right). The bottom panel shows a visually unremarkable ECG for Patient B with high probability A-ECG score for coronary artery disease (99%), and cardiac computed tomography angiography for same patient showing non-calcified plaque in mid right coronary artery with 75-99% stenosis and other foci of calcified plaque. An A-ECG score >50% is considered a high probability for significant coronary artery disease. ECG=electrocardiogram; A-ECG=advanced electrocardiography

The two A-ECG measures with the strongest association to significant CAD in our study were pertaining to QRS axis direction and vector magnitude. QRS axis shifts in the ECG occur with advancing age and have been associated with high-grade coronary stenosis in functional testing as well as higher long-term mortality in acute coronary syndrome (ACS) populations(17–19). Lower QRS voltage is seen in patients with multi-vessel coronary artery disease, as well the prediction of increased mortality risk in infiltrative cardiomyopathies such as amyloidosis (20, 21). We included increased R-wave duration in our A-ECG score – a contributor to overall QRS duration reflecting delayed intraventricular conduction. Studies have shown QRS prolongation as an independent predictor of cardiovascular outcomes in patients with CAD and ACS (22, 23). Lastly, we posit that the decreased R-wave amplitude in our significant CAD group may reflect overall reduced QRS vector magnitude discussed earlier. The localisation to lead aVF in our study may suggest a higher prevalence of inferior wall ischaemia or infarction in our cohort, with derived VCG patterns previously shown to have increased sensitivity compared to ECG for the identification of myocardial infarction in this territory(24, 25).

Using similar A-ECG analysis methods described in prior studies, we optimised a four-parameter A-ECG score demonstrating high negative predictive value and good sensitivity and specificity in excluding significant CAD as a cause for symptoms. Importantly, the calculated inverse negative likelihood ratio indicates a 6-fold increase in probability of absence of significant CAD, independent of disease prevalence. The A-ECG test has the potential to be used as a screening tool in the RACPC setting, though further prospective clinical studies are needed.

## Limitations

A limitation to this study is the relatively low proportion of patients with significant CAD, however this is to be expected from a low-intermediate risk chest pain population. An A-ECG score derived with more variables would require a very large sample size in order to identify a large absolute number of patients with significant disease. Although cross-validation of the A-ECG score was performed using bootstrapped resampling across the entire dataset, reassessment in a separate validation cohort would improve diagnostic confidence further.

Our study was performed retrospectively and without a control group of healthy volunteers. Given all patients presented with symptoms concerning for cardiac ischaemia, it is certainly possible that other diagnoses may be present and confounding, such as coronary microvascular dysfunction.

## Conclusions

A-ECG can rule out significant CAD on CCT with a high negative predictive value and good sensitivity and specificity using four variables derived from the 12-lead ECG. This supports the utility of A-ECG in the chest pain clinic setting to assist in risk stratification and identifying patients who would benefit from further testing or not. However, further large scale prospective trials are needed to validate this.

## Data Availability

All data produced in the present work are contained in the manuscript.

## References

1. Skinner JS, Smeeth L, Kendall JM, Adams PC, Timmis A. NICE guidance. Chest pain of recent onset: assessment and diagnosis of recent onset chest pain or discomfort of suspected cardiac origin. Heart (British Cardiac Society). 2010;96(12):974–8.

2. Yu C, Sheriff J, Ng A, Brazete S, Gullick J, Brieger D, et al. A Rapid Access Chest Pain Clinic (RACPC): Initial Australian Experience. Heart, lung & circulation. 2018;27(11):1376–80.

3. Black JA, Cheng K, Flood JA, Hamilton G, Parker S, Enayati A, et al. Evaluating the benefits of a rapid access chest pain clinic in Australia. Medical journal of Australia. 2019;210(7):321–5.

4. Backus BE, Six AJ, Kelder JC, Bosschaert MAR, Mast EG, Mosterd A, et al. A prospective validation of the HEART score for chest pain patients at the emergency department. International journal of cardiology. 2013;168(3):2153–8.

5. Kozor R, Mooney J, Lowe H, Kritharides L, Altman M, Klimis H, et al. Rapid Access Chest Pain Clinics: An Australian Cost-Benefit Study. Heart, lung & circulation. 2022;31(2):177–82.

6. Sox HCJ, Garber AM, Littenberg B. The resting electrocardiogram as a screening test: a clinical analysis. Annals of internal medicine. 1989;111(6):489–502.

7. Ashley EA, Raxwal V, Froelicher V. An evidence-based review of the resting electrocardiogram as a screening technique for heart disease. Progress in cardiovascular diseases. 2001;44(1):55–67.

8. Schlegel TT, Kulecz WB, Feiveson AH, Greco EC, DePalma JL, Starc V, et al. Accuracy of advanced versus strictly conventional 12-lead ECG for detection and screening of coronary artery disease, left ventricular hypertrophy and left ventricular systolic dysfunction. BMC cardiovascular disorders. 2010;10(1):28-.

9. Zong Y, Maanja M, Chaireti R, Schlegel TT, Ugander M, Antovic JP. Substantial prevalence of subclinical cardiovascular diseases in patients with hemophilia A evaluated by advanced electrocardiography. Journal of electrocardiology. 2020;58:171–5.

10. Sapra R, Hallqvist L, Schlegel TT, Ugander M, Bell M, Maanja M. Predicting peri- operative troponin elevation by advanced electrocardiography. Journal of electrocardiology. 2021;68:1–5.

11. Maanja M, Wieslander B, Schlegel TT, Bacharova L, Abu Daya H, Fridman Y, et al. Diffuse Myocardial Fibrosis Reduces Electrocardiographic Voltage Measures of Left Ventricular Hypertrophy Independent of Left Ventricular Mass. Journal of the American Heart Association. 2017;6(1):n/a.

12. Maanja M, Schlegel TT, Kozor R, Lundin M, Wieslander B, Wong TC, et al. The electrical determinants of increased wall thickness and mass in left ventricular hypertrophy. Journal of electrocardiology. 2020;58:80–6.

13. Excellence NIfHaC. Clinical guideline CG95 “Chest pain of recent onset: assessment and diagnosis.”. NICE. London, England 2010.

14. Rochitte CE, Gottlieb I, de Roos A, Cox C, Bush DE, Hoe J, et al. Diagnostic performance of coronary angiography by 64-row CT angiography. The New England journal of medicine. 2008;359(22):2324.

15. Mowatt G, Cummins E, Waugh N, Walker S, Cook J, Jia X, et al. Systematic review of the clinical effectiveness and cost-effectiveness of 64-slice or higher computed tomography angiography as an alternative to invasive coronary angiography in the investigation of coronary artery disease. Health technology assessment (Winchester, England). 2008;12(17):iii–iv.

16. Six AJ, Cullen L, Backus BE, Greenslade J, Parsonage W, Aldous S, et al. The HEART score for the assessment of patients with chest pain in the emergency department: a multinational validation study. Critical pathways in cardiology. 2013;12(3):121–6.

17. Punkka O, Kurvinen H-J, Koivula K, Eskola MJ, Martiskainen M, Huhtala H, et al. The prognostic significance of the electrical QRS axis on long-term mortality in acute coronary syndrome patients - The TACOS study. Journal of electrocardiology. 2022;73:22–8.

18. Shiran A, Halon DA, Merdler A, Makhoul N, Khader N, Ben-David J, et al. Accuracy of Exercise-Induced Left Axis QRS Deviation as a Specific Marker of Left Anterior Descending Coronary Artery Disease. Cardiology. 1998;89(4):297–302.

19. Lindow T, Palencia-Lamela I, Schlegel TT, Ugander M. Heart age estimated using explainable advanced electrocardiography. Scientific reports. 2022;12(1):9840-.

20. Kobayashi AMD, Misumida NMD, Aoi SMD, Kanei YMD. Low QRS Voltage on Presenting Electrocardiogram Predicts Multi-vessel Disease in Anterior ST-segment Elevation Myocardial Infarction. Journal of electrocardiology. 2017;50(6):870–5.

21. Saad JM, Ahmed AI, Alahdab F, Rifai MA, Han Y, Alfawara MS, et al. PROGNOSTIC VALUE OF LOW QRS VOLTAGE IN PATIENTS WITH TRANSTHYRETIN CARDIAC AMYLOIDOSIS. Journal of the American College of Cardiology. 2023;81(8):732-.

22. Schinkel AFLMD, Elhendy AMD, van Domburg RTP, Biagini EMD, Rizzello VMD, Veltman CEMD, et al. Prognostic Significance of QRS Duration in Patients With Suspected Coronary Artery Disease Referred for Noninvasive Evaluation of Myocardial Ischemia. The American journal of cardiology. 2009;104(11):1490–3.

23. Baslaib FMD, Alkaabi SMD, Yan ATMD, Yan RTMD, Dorian PMD, Nanthakumar KMD, et al. QRS prolongation in patients with acute coronary syndromes. The American heart journal. 2010;159(4):593–8.

24. Hurd Ii HP, Starling MR, Crawford MH, Dlabal PW, O’Rourke RA. Comparative accuracy of electrocardiographic and vectorcardiographic criteria for inferior myocardial infarction. Circulation (New York, NY). 1981;63(5):1025–9.

25. Starr JW, Wagner GS, Behar VS, Walston Iind A, Greenfield Jr JC. Vectorcardiographic criteria for the diagnosis of inferior myocardial infarction. Circulation (New York, NY). 1974;49(5):829–36.

